# Social Media Insights Into US Mental Health Amid the COVID-19 Pandemic. A Longitudinal Twitter Analysis (JANUARY-APRIL 2020)

**DOI:** 10.1101/2020.12.01.20241943

**Authors:** Danny Valdez, Marijn ten Thij, Krishna Bathina, Lauren Alexandra Rutter, Johan Bollen

## Abstract

**Background:** The COVID-19 pandemic led to unprecedented mitigation efforts that disrupted the daily lives of millions. Beyond the general health repercussions of the pandemic itself, these measures also present a significant challenge to the world’s mental health and healthcare systems. Considering traditional survey methods are time-consuming and expensive, we need timely and proactive data sources to respond to the rapidly evolving effects of health policy on our population’s mental health. Significant pluralities of the US population now use social media platforms, such as Twitter, to express the most minute details of their daily lives and social relations. This behavior is expected to increase during the COVID-19 pandemic, rendering social media data a rich field from which to understand personal wellbeing.

**Purpose:** Broadly, this study answers three research questions: RQ1: What themes emerge from a corpus of US tweets about COVID-19?; RQ2: To what extent does social media use increase during the onset of the COVID-19 pandemic?; and RQ3: Does sentiment change in response to the COVID-19 pandemic?

**Methods:** We analyzed 86,581,237 public domain English-language US tweets collected from an open-access public repository in three steps^1^. First, we characterized the evolution of hashtags over time using Latent Dirichlet Allocation (LDA) topic modeling. Second, we increased the granularity of this analysis by downloading Twitter timelines of a large cohort of individuals (n = 354,738) in 20 major US cities to assess changes in social media use. Finally, using this timeline data, we examined collective shifts in public mood in relation to evolving pandemic news cycles by analyzing the average daily sentiment of all timeline tweets with the Valence Aware Dictionary and sEntiment Reasoner (VADER) sentiment tool^2^.

**Results:** LDA topics generated in the early months of the dataset corresponded to major COVID-19 specific events. However, as state and municipal governments began issuing stay-at-home orders, latent themes shifted towards US-related lifestyle changes rather than global pandemic-related events. Social media volume also increased significantly, peaking during stay-at-home mandates. Finally, VADER sentiment analysis sentiment scores of user timelines were initially high and stable, but decreased significantly, and continuously, by late March.

**Discussion & Conclusion:** Our findings underscore the negative effects of the pandemic on overall population sentiment. Increased usage rates suggest that, for some, social media may be a coping mechanism to combat feelings of isolation related to long-term social distancing. However, in light of the documented negative effect of heavy social media usage on mental health, for many social media may further exacerbate negative feelings in the long-term. Thus, considering the overburdened US mental healthcare structure, these findings have important implications for ongoing mitigation efforts.

## 1 Introduction

Beyond the obvious physical health ramifications of the COVID-19 pandemic, public health, and the greater medical community, is also bracing for a mental health crisis^3^. Within the span of four months, 45% of Americans indicated that the COVID-19 pandemic had taken a toll on their mental health, reporting higher levels of sadness and worsening of chronic psychiatric conditions^4^. Yet, despite an abundance of anecdotal evidence and peer-reviewed editorials that identify the potential mental-health fallout of this public health crisis, the extent of these effects is empirically unknown.

Scientists mobilized quickly to measure many facets of the pandemic, including potential mental-health effects. However, the time-consuming and costly nature of survey development^5^ and instrument validation make it difficult to draw real-time conclusions^6^, especially amidst rapidly evolving news cycles that shift pandemic related discourse. In the absence of survey data, social media represents a potentially valuable data source for studying emergent social issues, including the effect of those issues on behaviors and social mood^7^. Repeated tracking of social media data can provide a diachronic perspective on public morale and collective changes in sentiment, as participants voluntarily contribute to narratives, providing unprompted and diverse understandings of various issues^8–10^. Numerous scholars have successfully used social media data to identify trends and nuances in public mood using a combination of machine learning and artificial intelligence approaches. Some examples include comparing the happiness of users to their online social networks^11,12^, identifying detailed predictors of mood through social media feeds^7^, predicting cognitive distortions expressed among groups at-risk of mental health disorders^13^, tracking the emotions of social media users at high resolution^14,15^, and mapping negative affectivity among users with internalizing disorders^16^. Collectively, these studies demonstrate the feasibility and value of using sentiment analysis on social media data to study societal mood and well-being, as well as biomedical signals among social media users that can provide useful proxies for mental health^13,17–19^. In fact, these approaches may be especially useful considering the speed with which the pandemic became an acute socio-economic phenomenon, the pervasiveness of COVID-19 related content available online, and the natural reaction of many to post on social media about pandemic-related events.

Indeed, throughout the COVID-19 pandemic, individuals have sought out crisis-related news at increased capacities^20^, leading to a collective increase in global social media use^21^. This renders social media data about the COVID-19 pandemic a powerful source of information from which to draw real-time conclusions about aggregate social well-being during an unprecedented public health event. However, we must remember that just as survey data are prone to biases, so are data derived from social media^22^. Therefore, to draw accurate inferences about mood/sentiment and mental health, we must remain cognizant of the type of analysis performed and what the analysis represents to measure nuanced aspects of sentiment. Here we contrast (1) topics discussed, (2) topic-related sentiment, and (3) personal sentiment to arrive at a more comprehensive and accurate assessment of changes in sentiments expressed through social media and their relevance to public health in the United States.

Broadly, this study answers three research questions: (1) RQ1: What themes emerge from a corpus of US tweets about COVID-19? (2) RQ2: To what extent does social media use increase during the onset of the COVID-19 pandemic? And (3) RQ3: What patterns emerge from longitudinal tracking of sentiment during the onset of the COVID-19 pandemic? To address these research questions, we analyzed a large-scale set of Twitter data that are strictly relevant to the topic of COVID-19 in the United States from January 22, 2020 onwards. Using this data, we also compiled a second corpus of individual, geolocated social media timeline data from the same period to understand changes in personal sentiment as a proxy for mental health and evolving US perceptions of the COVID-19 pandemic.

## 2 Methods

### 2.1 Data

We collected two distinct data sets, each reflecting different aspects of changes in social media behavior before and during the COVID-19 pandemic. The first dataset of tweets, collected from an open-access repository containing all COVID related tweets published in the United States^1^, was designed to capture topical differences (i.e. themes) in the Twitter discussion during the events that marked the onset of the pandemic. The repository provides a list of Tweet IDs, which we used to extract tweet content from Twitter’s API (endpoint: GET statuses/show/id). We downloaded each tweet as well as the standard meta-data provided by Twitter. Specifically, we retrieved COVID-19 related tweets posted between January 22, 2020 (first day of data collection and roughly one week prior to the first confirmed US COVID-19 case) through April 9, 2020 (the middle of social distancing efforts). Hereafter, we refer to this set of tweets as the “COVID-19 corpus”, (n = 86,581,237 tweets). Please refer to Figure 1 for a visual representation of this dataset and how it was retrieved.

**Figure 1.**
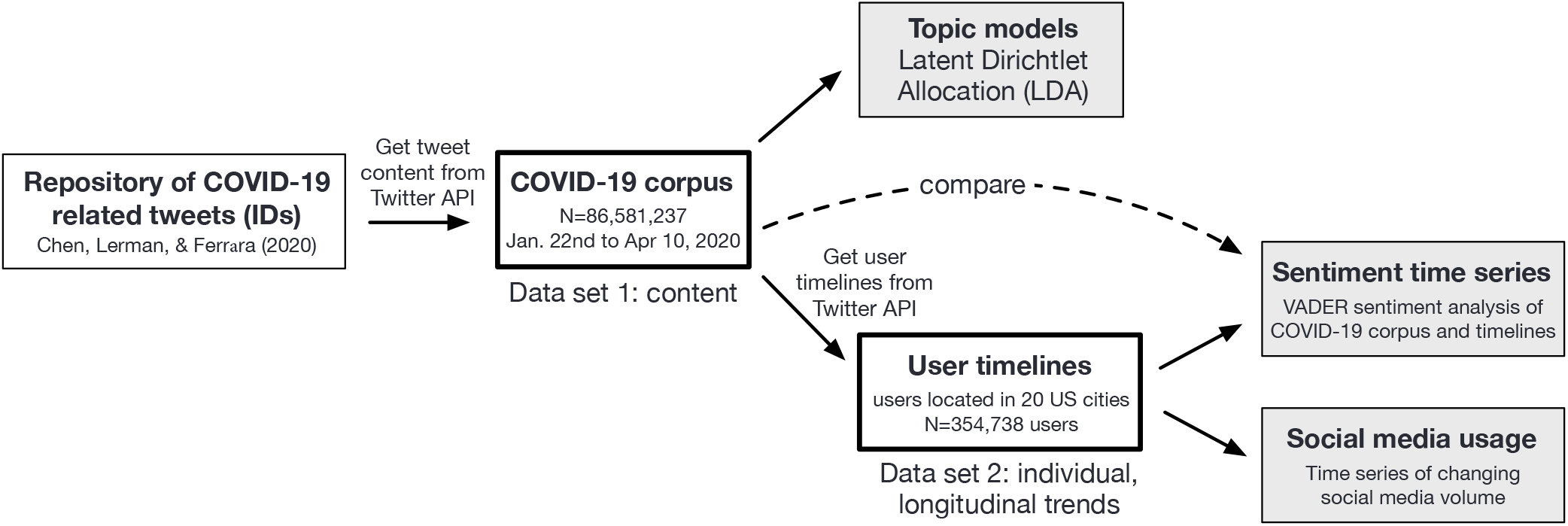
This diagram illustrates the procedure undertaken to procure Tweet IDs from an open-access COVID-19 repository. These tweet IDs were run through Twitter’s API to create two distinct data sets. The COVID-19 corpus, which contains all COVID-19 related English-language tweets published in the United States between January-April 2020. The User time-line data contains the 3200 most recent tweets of users residing in the 20 cities most-affected by COVID-19.

Second, to gauge fluctuations of personal activity and mood at the individual, rather than topical, level, we downloaded the twitter timelines (i.e. the 3,200 most recent tweets) of individual social media users who contributed to the COVID-19 corpus and also resided within the 20 US cities with the most COVID-19 cases per 100,000 people from the Twitter API (endpoint: GET statuses/user_timeline). These timelines capture the changing behavior and emotions of individual Twitter users during the COVID-19 pandemic, but do not strictly pertain to tweets exclusively related to COVID-19. We refer to this data as user-timeline data; n = 354,738 users, (n = 69,349,479 tweets), as shown in Figure 1. All tweets in either dataset were scrubbed of any personally identifiable information in accordance with ethical social media use practices.

To ensure that we are measuring expressed sentiment in our data we exclude non-English tweets and, within the user-timeline data, specifically, retweets, and biasing key-words, including: (1) “coronavirus”, (2) “COVID-19”, (3) “pandemic”, among others. These words were removed because they inherently carry a negative connotation and their inclusion would artificially decrease sentiment given that the corpus itself is composed of COVID-19 related content. In other words, because users are naturally tweeting about “coronavirus”, “virus” and the “pandemic”, the inclusion of those words may not necessarily reflect the individuals’ well-being. Note, the resulting sample sizes for each corpora exceeds the mean observed in a recent scoping survey of the literature on social media analytics for public health by several orders of magnitude (n = 20,000)^23^, resulting in ample representation with which to conduct our analyses. Additionally, previous studies have used large-scale sentiment analysis to accurately predict social mood^24^, and how sentiment expressed on social media correlates with psychological well-being^25^. Thus, the use of sentiment analysis for this study is appropriate.

### 2.2 Analyses

#### LDA Topic Models

Latent Dirichlet Allocation (LDA) topic models are unsupervised machine-learning tools that perform probabilistic inferences to consolidate large volumes of text data into manageable themes^26^. Simply, words with high probabilities of association (i.e. a high likelihood to appear in proximity with other words) are grouped together to form a latent theme, or topic, that qualitatively represents a content area within the collection of text. These methods have been applied in myriad ways, e.g. to determine common themes in product reviews^27^, to map themes within bodies of scientific literature^28^, and to identify themes in social media data^29^. Thus, these tools are appropriate for exploratory analyses that seek to consolidate dense text data.

#### Sentiment Analysis

Sentiment analysis refers to a set of supervised and/or unsupervised Machine Learning (ML) and Natural Language Processing (NLP) techniques that extract affective or emotional indicators from text, e.g. to determine whether a tweet expresses a negative or positive emotion about policy^24^. Here we used the Valence Aware Dictionary and sEntiment [sic] Reasoner (VADER)^2^ to gauge the emotional valence of tweets. VADER is a rule-based, open-source tool that recognizes common terms, idioms, abbreviations, and jargon, while accounting for grammatical structures, such as punctuation, negation, hedging, and magnification, that are commonly employed in the vernacular of social media platforms. The VADER lexicon is one of the largest of its kind containing over 7,500 common terms that are each rated for their emotional valence by 10 independent human raters. However, the word “virus” and its many variations (e.g. “viruses”, “viral”) are not part of the VADER lexicon, meaning changes in the frequency of these words will not bias VADER scores. VADER has been extensively validated for Twitter content^30^, showing some of the highest accuracy and coverage for tweets in a benchmark of more than 20 sentiment analysis tools^31^.

#### Change-point detection

We applied the Pruned Exact Linear Time (PELT) change-point detection algorithm to identify significant changes in tweet volume and sentiment^32^. Change-point detection algorithms perform a set of mathematical operations over a time series (a series of time-based observations) to identify points in time at which the statistical properties of the time-series data change significantly^33^. The PELT algorithm specifically attempts to find a set of change points for a given time series, such that their number and location in time minimizes a given segmentation cost. We chose the PELT algorithm over other similar change detection algorithms because it is considered to be a more conservative estimate (preferring not to identify change points unless strict conditions are satisfied), thus yielding more accurate detection of statistical changes^34^. Additionally, PELT uses an off-line approach to change detection^34^, meaning it can consider all possible data points when identifying significant changes, regardless of the type of data, while maintaining high levels of performance.

### 2.3 Procedure

RQ1: What themes emerge from a corpus of US tweets about COVID-19? We divided the COVID-19 corpus into daily segments and generated one topic model per day consisting of 20 topics each. We chose this number to reflect the widest possible span of themes while summarizing the major themes of the online discussion, which is a process used in previous studies^29,35^. We then looked at the top 20 associated words per topic and collapsed similar words into general themes, taking into account similarities of words (i.e. United States and US), and potential misspellings, which are common in social media posts. As an example, “hubei” and “wuhan” were collapsed into the theme “China”. We then found the frequency ratio (i.e. the number of occurrences of a certain word divided by the total number of words) of COVID-19 related themes (“China”, “US”, “pandemic”, “social distancing”, “Trump”, “home”, “lockdown” and “deaths”) and plotted them on a daily basis to show the evolution of topics over time, indicating both the contribution of the theme to all content of that day and the relative ranking of these terms amongst these themes. We used inter-coder agreement methods to arrive at mutually agreeable interpretations of collapsed themes^36^. RQ2: To what extent does social media use increase during the onset of the COVID-19 pandemic? For this analysis we used the user-timeline data instead of the general COVID-19 corpus (See Figure 1) because within-subject individual posting frequency is a better marker for tracking changes in social media use behavior^37^. Since Twitter’s API limits us to only the 3200 most recent postings per individual, we only selected individuals who posted on Twitter before January 22, 2020, retaining 354,738 users. This ensures that our analytic sample captures individual behavior in the 20-most affected US cities throughout the interval of interest (January 22, 2020 to April 9, 2020). We performed a seasonal decomposition— a method that separates the baseline, trend, and seasonal components of a time series— to determine whether we can observe increased Twitter use during the pandemic relative to events just prior. We then detected significant points of change using the PELT algorithm^38^. RQ3: What patterns emerge from longitudinal tracking of sentiment during the onset of COVID-19 pandemic? Sentiment can change because individuals discuss different topics, e.g. using pejorative terms such as “virus” more frequently, or because of personal, individual changes in how people actually feel. We therefore compared daily VADER sentiment scores for (1) the COVID-19 corpus (to gauge topical sentiment) and (2) user-timeline within-subjects data (to assess personal changes in sentiment) from January 22, 2020 (the first official day of data collection), through April 9, 2020. We determine change-points in the time-series of daily averaged VADER sentiment with the PELT detection algorithm so we can identify significant changes in sentiment throughout this time period.

## 3 Results

### RQ1: What themes emerge from a corpus of US tweets about COVID-19?

We consolidated the COVID-19 corpus (n= 86,581,237 tweets) into themes using LDA topic models. Figure 2 highlights the eight most salient topics and how their prominence (width of bars and rank) changed over time relative to major COVID-19 related milestones identified by the WHO.

**Figure 2.**
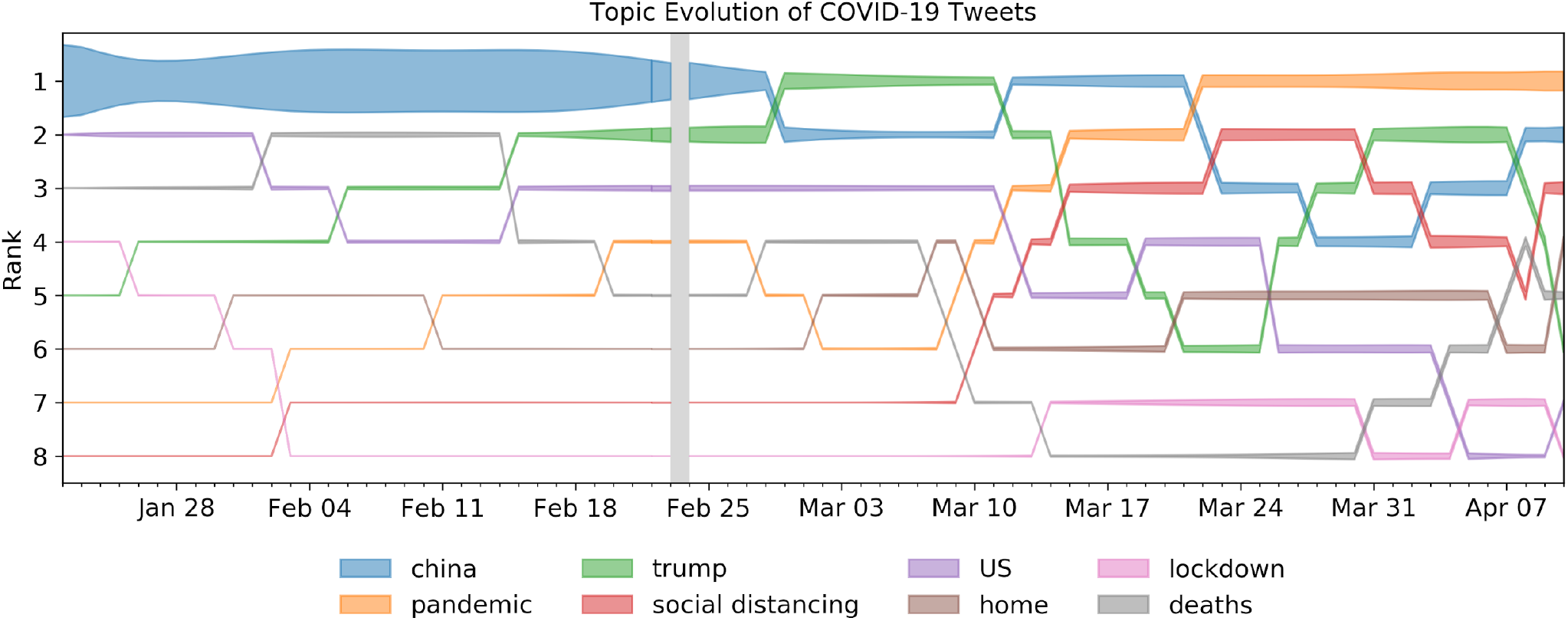
Topic-group rankings over time in the COVID-19 corpus. Each topic is ranked by its frequency ratio. The width of the bars indicates the fraction of words in the topic on a given day. The colors of the areas indicate which theme the corresponding area belongs to.

As shown in Figure 2, topics continued to rise and fall in prominence relative to emerging news cycles throughout the time period under consideration. Indeed, the majority of COVID-19 related Twitter activity focused on China in February. However, from March through April, as the novel coronavirus increasingly began to affect the US population, “China” became less prominent as more US-centered topics such as “lockdown” and “social distancing” emerged. Although China remained a prominent theme throughout the duration of interest, US-centered topics gradually came to dominate social media spaces.

### RQ2: To what extent does social media use increase during the onset of the COVID-19 pandemic?

Using user-timeline data, we compared the frequency of social media posts before and during the pandemic of 354,738 users individuals in 20 of the most-affected US cities (n= 69,349,479 tweets). Figure 3 highlights changes in posting volume between January 22, 2020 and April 9, 2020. The peaks and troughs (dashed line) of this graph show how seasonal and weekly cycles shape tweet volume. The solid line plots the trend in the time-series after removing cycles and seasonal effects (through seasonal decomposition), and the progressively darker shades of brown denote the number of cities that imposed mandatory lockdowns.

**Figure 3.**
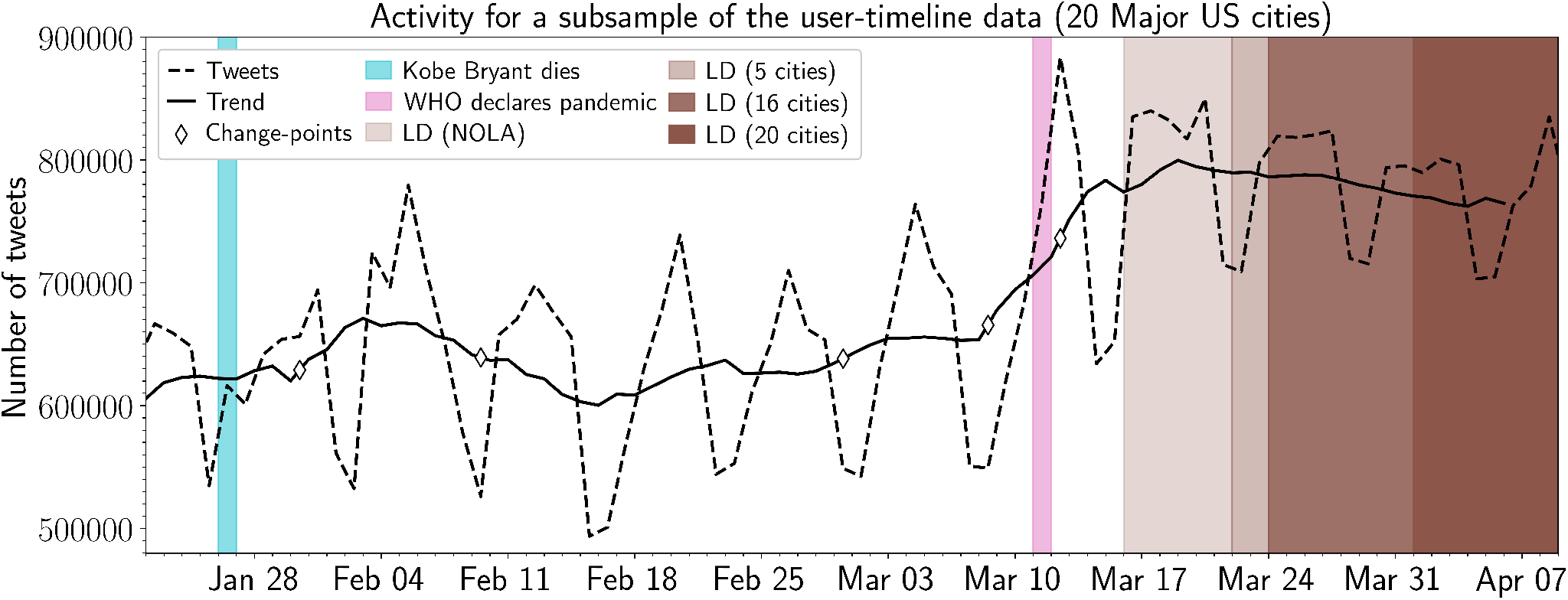
Number of daily tweets in a subsample of the user-timeline data (n=292,000). The dashed line displays the daily number of tweets in the subsample of the user-timeline data. The solid line indicates the underlying trend in the daily number of tweets (after removing its seasonal effects). The diamond markers indicate significant change-points indicated by the PELT-algorithm. The light-blue and pink annotations denote the death of Kobe Bryant (Jan 26, 2020) and the WHO declared COVID-19 as a global pandemic (March 11, 2020), respectively. The brown annotations indicate the dates at which lockdowns were enforced in the 20 considered cities (ranging March 16, 2020 to April 1, 2020). The opacity of the brown annotation indicates how many cities had enforced a lockdown at that date.

Generally, we observed a consistent upward trend in total Twitter volume from early to late March. The PELT change-point algorithm identified two significant volume changes on March 8 and 12 — around the time COVID-19 was declared a global pandemic (March 11) and President Trump’s national emergency declaration in the United States (March 14). The upward trend stabilized thereafter, albeit at higher observed volumes than prior to the onset of the COVID-19 pandemic. This supports the notion that individuals in our sample were more engaged with social media and made more use of it, possibly to discuss or obtain further information relevant to the news cycle.

### RQ3: What patterns emerge from longitudinal tracking of sentiment during the onset of the COVID-19 pandemic?

We applied the VADER sentiment tool to (1) the COVID-19 corpus (to assess sentiment of all US tweets about COVID-19; and (2) the user-timeline data (to track changes in user sentiment using their most recent 3,200 tweets). Figure 4 tracks sentiment relative to major COVID-19 related milestones for both datasets, with the orange line tracking the COVID-19 corpus, and blue line tracking the user-timeline data, respectively. In the COVID-19 corpus, there was an unmistakable increase in sentiment with two PELT-identified significant changes on March 9 (just before WHO classified COVID-19 as a pandemic) and March 19 (shortly after President Donald Trump declared a national emergency). Figure 5 further shows the percentage of positively scored COVID-19 tweets increased over time, reinforcing the positive trend.

**Figure 4.**
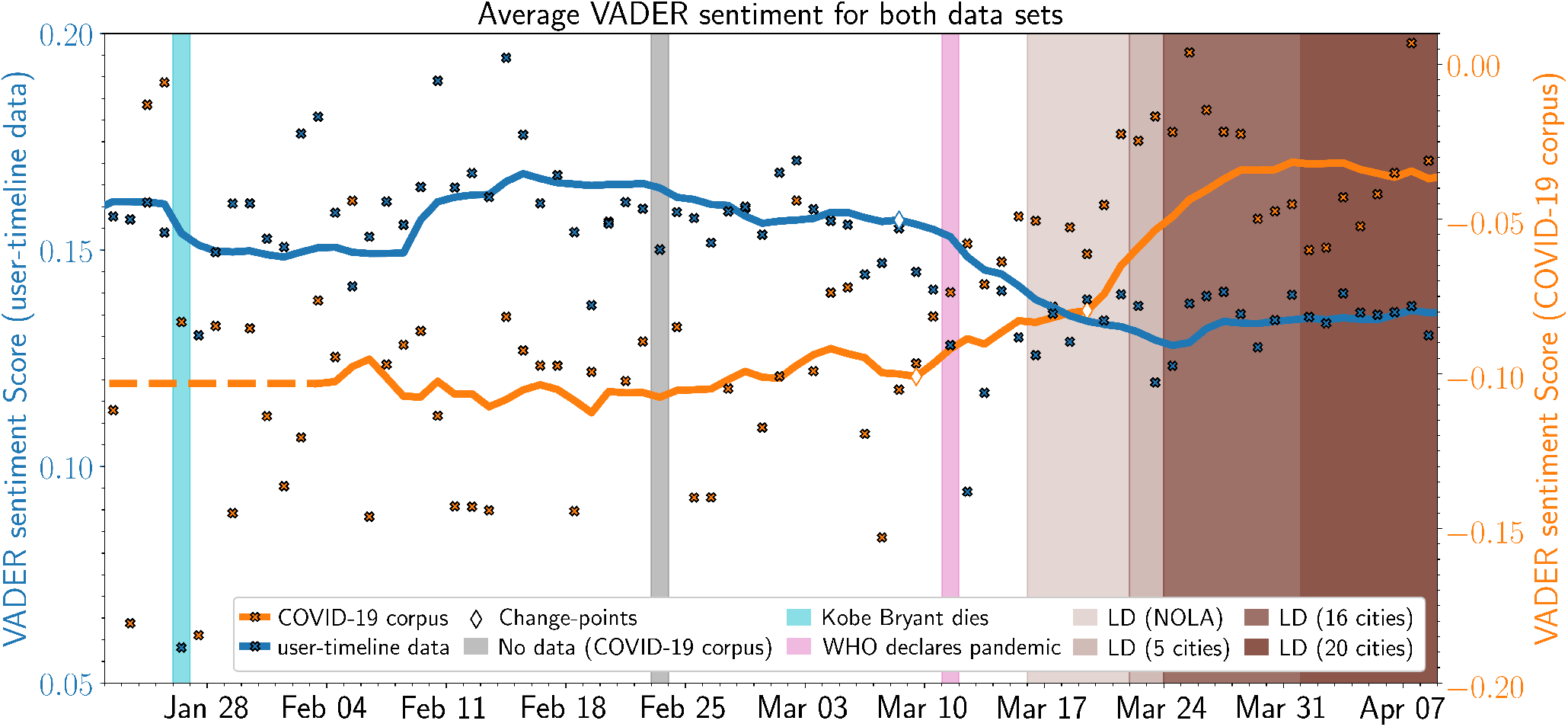
Distribution of non-zero VADER sentiment in the COVID-19 corpus & user-timeline data. The solid lines display the 14-day moving average and the crosses display the actual values of the daily average VADER sentiment (blue for the user-timeline data and orange for the COVID-19 corpus, respectively). The diamond markers indicate change-points indicated by the PELT-algorithm, the color of the edge of the diamonds refers to the time-series to which this change-point belongs. The light-blue, gray, and pink annotations denote the days of the death of Kobe Bryant (Jan 26, 2020), the day of missing data in the COVID-19 corpus (February 23, 2020), and the WHO declaring COVID-19 a global pandemic (March 11, 2020), respectively. The brown annotations indicate the dates at which lockdowns were enforced in the 20 considered cities (ranging March 16, 2020 to April 1, 2020). The opacity of the brown annotation indicates how many cities had enforced a lockdown at that date.

**Figure 5.**
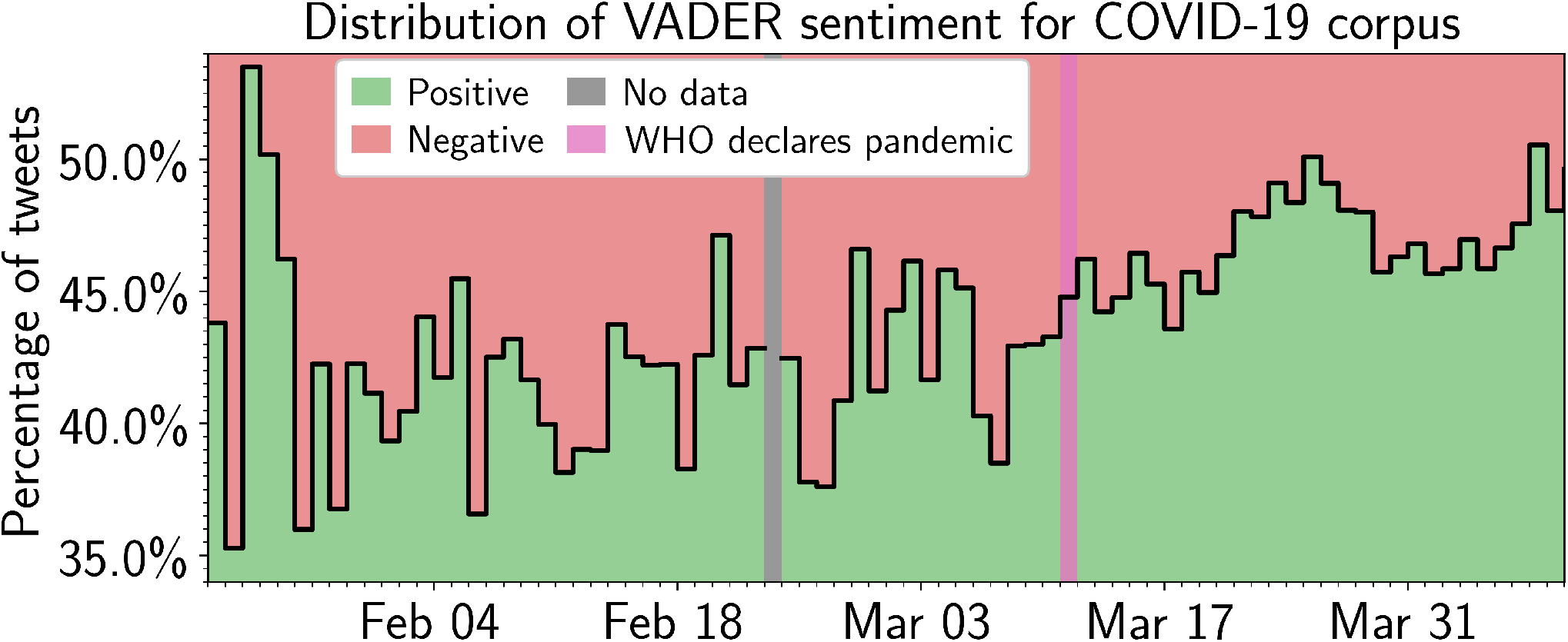
Distribution of positive and negative VADER sentiment in COVID-19 corpus. The solid line displays the fraction of tweets that hold positive or negative VADER sentiment in the user-timeline data (colored green for positive and red for negative sentiment). The gray and pink annotations denote the day of missing data (February 23, 2020) and the WHO declared COVID-19 as a global pandemic (March 11, 2020), respectively.

Conversely, the user-timeline data (which again contained the most recent 3,200 tweets of a given user) shows decreases in sentiment over the same period. The user-timeline data has one PELT-identified significant change a pandemic). Of note, there was a notable but short-lived drop in user before the WHO point on March 9 (just classified COVID-19 as sentiment on January 28– the day NBA player Kobe Bryant died in a helicopter crash.

## 4 Discussion

The purpose of this study was to draw conclusions about US mental health amid the COVID-19 pandemic using computational social media analytics of a publicly available repository containing all COVID-19 related tweets. Using the COVID-19 tweet repository released by Chen and colleagues, we expanded on their original topical analysis of COVID-19 content by examining both topics and sentiment specific to the US population to understand how the COVID-19 pandemic may be impacting social well-being. We used social media data as the medium for analysis for several reasons, in particular its usefulness to gauge real-time changes in mental health/social well-being before, during, and after rapid socio-economic changes^17^. Below we discuss the findings of this study in detail and highlight public health implications about social well-being during the onset of the COVID-19 pandemic.

### 4.1 COVID-19 Social Media Themes: Content is Reactionary to News Cycles

Bento and colleagues predicted that crisis-related information seeking would increase during the COVID-19 pandemic^20^. Numerous studies have supported that prediction by identifying increased panic within social media spaces as users react to COVID-19 news-related content on their feeds^39,40^. Through our LDA topic modeling analysis, we echo much of those findings as well; the topics uncovered by their LDA frequently correspond to the life cycles of COVID-19 related news and current events. For example, the name used to identify the virus on social media evolved to match changes in viral terminology as presented on news outlets– e.g. the novel coronavirus became coronavirus, COVID-19, and COVID-19 pandemic, respectively. The locations that emerged within the topics also corresponded to the movement of COVID-19 from mainland China to the United States. For example, in late January, “China” was a dominant theme in Twitter content (shown by the width of the bars in Figure 2) indicating the US public may have considered the then-classified epidemic a foreign matter (e.g. TWEET: “OMG China just shut down trading… Still think it’s just a flu?”). However, in later weeks/months, US-centered topics/tweets replaced “China” in prominence as more Americans became preoccupied with pandemic control measures at home (e.g. TWEET: “lol it’s wild seeing every single other country grappling with this virus in ways we KNOW the US will never do”). These findings suggest that as the COVID-19 pandemic became more prominent in the US, social media content changed to topics of more direct impact to the US, and with greater frequency of use (evidenced by a PELT-identified significant increase in Twitter volume during that time). This volume increase remained consistently above levels observed prior to the COVID-19 pandemic, which is possibly explained by several factors, including increased anxiety as the COVID-19 pandemic reached the United States, the isolating effects of state-wide stay at home orders, or other social fallouts driven by the pandemic. These findings collectively support those of Castillo and colleagues^41^, who contend that social media content follows the life-cycle of news stories. Thus, news, as a vehicle for crisis-related social media communication, should be studied more intently.

### 4.2 COVID-19 Topical Sentiment: Increased sentiment may indicate a priming effect

Given the expected negative physical and emotional outcomes of a major pandemic, we were surprised to observe that the sentiment of the COVID-19 corpus trended positively. However, this increase in sentiment is likely not indicative of actual changes in population sentiment, but rather the effect of a common topical bias in social media analytics^42^. Our COVID-19 data set was specifically selected to contain tweets that related to the topic of COVID-19. Hence, its sentiment scores will reflect the language used to discuss this particular topic, and not necessarily the underlying emotion of the population. Topically selected social media posts are likely primed by news cycles^43^ and shows of false optimism—a phenomena where individuals tend to post content that is more positive or optimistic than their true emotions^44^. Previous research has furthermore shown that language in general is biased towards positivity, especially when posts are collected for a random topic^30^. For example, language used to compose a post about COVID-19 may contain trending verbiage or framing devices, including showcasing support of groups frequently part of news cycles (e.g. TWEET: “Great news…it’s a welcome burden lifted off our incredible nurses and medical first responders”). Similarly, users may just be conveying positivity through carefully selected ‘popular’ words around a trending topic (e.g. TWEET: “No matter how hard the situation nowadays during the pandemic outbreak, we should keep being positive and optimistic”). Within the VADER lexicon, many of the words commonly used in this context are scored positively (great, welcome, incredible, positive, optimistic), which also artificially inflates sentiment ratings. Thus, we posit that topically driven tweet samples may not validly reflect actual changes in population mood, but rather topic-driven language sentiment. This justifies our approach to analyze within-subject timelines of individual posts which are not necessarily bound by the criterion of strictly being COVID-19 related, thereby increasing the odds of reflecting personal changes in mood trajectories.

### 4.3 User-timeline sentiment: Lower timeline sentiment may indicate decreased social well-being

By contrast, we found a negative trajectory in sentiment scores for the user-timeline data. This means that, while content in the COVID-19 corpus trended positively (possibly due to priming), relative to the totality of their timelines, our sample mood is lower than it once was. Thus, this comparison gives us deeper insight into underlying mood and sentiment. This finding further supports the assertion that the positivity conveyed in social media posts may not validly reflect what people are feeling at a given moment. Indeed, that positivity may be serving as a veneer posted “in the moment” to convey positivity during a time of uncertainty. To obtain an accurate assessment of mood and well-being, other reference points (in this case social media posts prior to the pandemic) are needed to examine in the moment sentiment relative to their prior histories. Within our timeline data, we captured one event that occurred prior to the pandemic that also affected sentiment scores relative to timeline histories—the passing of NBA basketball player Kobe Bryant. The effect of Bryant’s passing led to a sharp decline in sentiment, which lasted approximately 24 hours before returning to levels observed previously. Regarding the pandemic, after the PELT-identified shift on March 8, sentiment scores (relative to timelines) not only became lower, but consistently lower, and did not return to levels observed before the pandemic reached the United States. This trend may hold implications into the longitudinal effects of the pandemic and subsequent impacts on US mood/social sentiment, as this continued trend may be indicative of a long-lasting shift in mood and well-being.

### 4.4 Social Media Use in Times of Crisis

Analyses of aggregated social media feeds are shown to adequately predict other phenomena, including the stock market^11^, political leanings^45^, and when analyzed through a time-series, collective shifts in general mood^46^. Our study contributes to this body of literature by highlighting the disparity between how Americans portray themselves on social media, versus the latent sentiment they may be experiencing during times of crisis. Generally, Americans were not posting social media content about COVID-19 prior to the first documented US case. Once COVID-19 became a reality in the United States, however, there was a continued increase in the total number of US-based tweets about the novel coronavirus, indicative of growing social media use in our sample. By analyzing the COVID-19 corpus and user-timeline data separately, but with the same analytic procedures, we see divergent findings that reinforce the difference between in-the-moment portrayals versus the longitudinal information that can be gleaned from individual timeline analyses. For example, tweets and posts about COVID-19 may attempt to be lighthearted or convey optimism; however, individually, social media users may not be as optimistic as they were prior to the pandemic. During an unprecedented public health crisis it is therefore important to look beyond the topical focus of messages on social media that reference the crisis itself as a proxy of public mood, as they are likely to be affected by other influences (e.g., political framing, projecting hope). Ultimately, our findings exemplify COVID-19 as a case-study in social media behavior, whose outcomes should be generalized to other crisis-related events.

## 5 Concluding Remarks and Implications

At the time of writing, the US COVID-19 death toll was just over 136,000, with millions of people disrupted by the various effects of the pandemic. This study elucidates the possible mental health effects of the COVID-19 pandemic among Twitter users using a computational approach to analyze a corpus of all archived, US-based COVID-19 tweets from January-April 2020. These analyses revealed, to varying extents, how the pervasiveness of COVID-19 content available on social media and abrupt shift in lifestyles may be negatively affecting social sentiment relative to points just prior to the pandemic. Given that sentiments expressed on social media have been used as a proxy for mental well-being^47^ these findings support calls from public health and medical scholars who contend that mental health is an urgent concern during the COVID-19 pandemic, especially as our findings illustrate a declining trend in sentiment. Thus, we encourage further research on US mental health status amid the pandemic using survey methods or other primary data collections to substantiate our findings with testable outcomes. We also call for more research on mental-health interventions amid the COVID-19 pandemic, with particular attention to modality (i.e. in-person vs virtual), and the efficacy of those efforts.

### 5.1 Limitations

Our study is subject to limitations. First, Twitter requires users to opt-in to geo-tagging features. Consequently, any information inferred about the user’s city of residence is often limited to self-reported data as specified on profile pages. This may mean that some of our timeline data may not originate from the city specified by a user, as this information can easily be misrepresented^48^ (i.e. stating they live in New York, NY, but actually residing in Newark, NJ). We also acknowledge a likely bias regarding key demographic information, including age, gender, and socioeconomic status among likely social media users^49^, in addition to the temporal, spatial, and geographic patterns that may affect how sentiment is expressed on social media (e.g. older adults posting early in the morning versus younger adults posting late at night; or urban-based versus rural users)^50^. It is also, as of yet, not possible to accurately diagnose someone with a mental health condition through social media feeds alone, although research has indicated social media content contains important signals with respect to mental health and biomedical signals. Thus, we relied on trend data to draw inferences about the possibility of mental health decline based on average sentiment scores^51^. However, these limitations do not diminish the importance or validity of this work. Rather, they create avenues for additional research that expand on the findings of this paper and leverage the limitations inherent to social media data– such as measuring cognitive distortions on Twitter based on posting time, or approaches that measure diagnosable mental health conditions through social media data, particularly during times of increased panic and crisis. And, because social media has been widely used to draw conclusions about public mood through large-scale sentiment analysis procedures^20,52^, we contend our approach is appropriate to draw the conclusions discussed herein.

## Data Availability

Data and source code are available by request.

